# Geographic Disparities and Predictors of COVID-19 Hospitalization Risk in the St. Louis Area, Missouri (USA)

**DOI:** 10.1101/2021.10.21.21265289

**Authors:** Morganne Igoe, Praachi Das, Suzanne Lenhart, Alun L. Lloyd, Lan Luong, Dajun Tian, Cristina Lanzas, Agricola Odoi

## Abstract

**Background:** COVID-19 has overwhelmed the US healthcare system, with over 44 million cases and over 700,000 deaths as of October 6, 2021. There is evidence that some communities are disproportionately affected. This may result in geographic disparities in COVID-19 hospitalization risk that, if identified, could guide control efforts. Therefore, the objective of this study is to investigate Zip Code Tabulation Area (ZCTA)-level geographic disparities and identify predictors of COVID-19 hospitalization risk in the St. Louis area.

**Methods:** Hospitalization data for COVID-19 and several chronic diseases were obtained from the Missouri Hospital Association. ZCTA-level data on socioeconomic and demographic factors were obtained from the US Census Bureau American Community Survey. Age-adjusted COVID-19 and several chronic disease hospitalization risks were calculated. Geographic disparities in distribution of COVID-19 age-adjusted hospitalization risk, socioeconomic and demographic factors as well as chronic disease risks were investigated using choropleth maps. Predictors of ZCTA-level COVID-19 hospitalization risks were investigated using global negative binomial and local geographically weighted negative binomial models.

**Results:** There were geographic disparities of COVID-19 hospitalization risks. COVID-19 hospitalization risks were significantly higher in ZCTAs with high diabetes hospitalization risks (p<0.0001), high risks of COVID-19 cases (p<0.0001), as well as high percentages of black population (p=0.0416) and populations with some college education (p=0.0005). The coefficients of the first three predictors varied across ZCTAs, implying that the associations between COVID-19 hospitalization risks and these predictors varied by geographic location. This implies that a “one-size-fits-all” approach may not be appropriate for management and control.

**Conclusions:** There is evidence of geographic disparities in COVID-19 hospitalization risks that are driven by differences in socioeconomic, demographic and health-related factors. The impacts of these factors vary by geographical location with some factors being more important predictors in some locales than others. Use of both global and local models leads to a better understanding of the determinants of geographic disparities in health outcomes and utilization of health services. These findings are useful for informing health planning to identify geographic areas likely to have high numbers of individuals needing hospitalization as well as guiding vaccination efforts.

## Background

There have been over 184 million confirmed Coronavirus Disease 2019 (COVID-19) cases and over 4 million deaths worldwide as of July 8, 2021, with over 33 million confirmed cases and over 600,000 deaths in the United States [1]. As of the same date, the state of Missouri has reported over 633,000 cases and 9,700+ deaths. The disease has overwhelmed many United States hospital systems, with large numbers of patients requiring critical care and interventions such as mechanical ventilation [2–4]. This surge of COVID-19 patients has put strain on hospital resources, potentially impacting the care available to both COVID-19 and non-COVID-19 patients [5].

There is evidence of geographic disparities in the severity of the disease with certain population groups experiencing more severe disease than others. These disparities might be driven by population characteristics such as socioeconomic, demographic, and chronic disease factors [6–10]. For instance, there is evidence that Non-Hispanic American Indian, non-Hispanic Black, and Hispanic people have higher hospitalization risks than their non-Hispanic Asian and non-Hispanic White counterparts [11].There are also reports that conditions such as diabetes mellitus, obesity, chronic lung conditions, renal disease, cancer, and cardiovascular disease may increase the severity of the condition and risk of hospitalizations among COVID-19 patients with these co-morbidities [6, 12–14]. Identifying geographic disparities in COVID-19 hospitalization risks and determinants of these disparities is important in providing information to guide hospital preparedness to handle the patient surge and for targeting resources for public health efforts to control the condition at the community level.

Improved understanding of the geographic disparities and predictors of COVID-19 hospitalization risk at the local level, such as the Zip Code Tabulation Areas (ZCTA), would help identify local geographic areas with higher needs for hospital beds and other healthcare resources. This is useful information for healthcare planning and service provision at the local level. It may also inform vaccination efforts by helping to identify areas where higher vaccination coverage may have the largest effect in reducing hospital burden. Therefore, the objective of this study is to identify geographic disparities and predictors of COVID-19 hospitalization risk in the St. Louis Area, Missouri, United States.

## Methods

### Study Area

This study was performed in an area of Missouri that includes 108 ZCTAs located in St. Charles county, St. Louis county, and St. Louis City as well as parts of Jefferson, Franklin and Warren counties (**Figure 1**). The area had a population of approximately 2 million people comprising 74% white, 20% black, 3% Hispanic/Latino, and 3% Asian. Forty-eight percent of the population was male, while 52% was female. Thirty-one percent of the population was older than 25 years old, 53% was 25-64, and 16% was older than 65 years of age. The ZCTA-level population density varied from 10 people per square mile in St. Charles County to 9,368 people per square mile in St. Louis City County.

**Figure 1:**
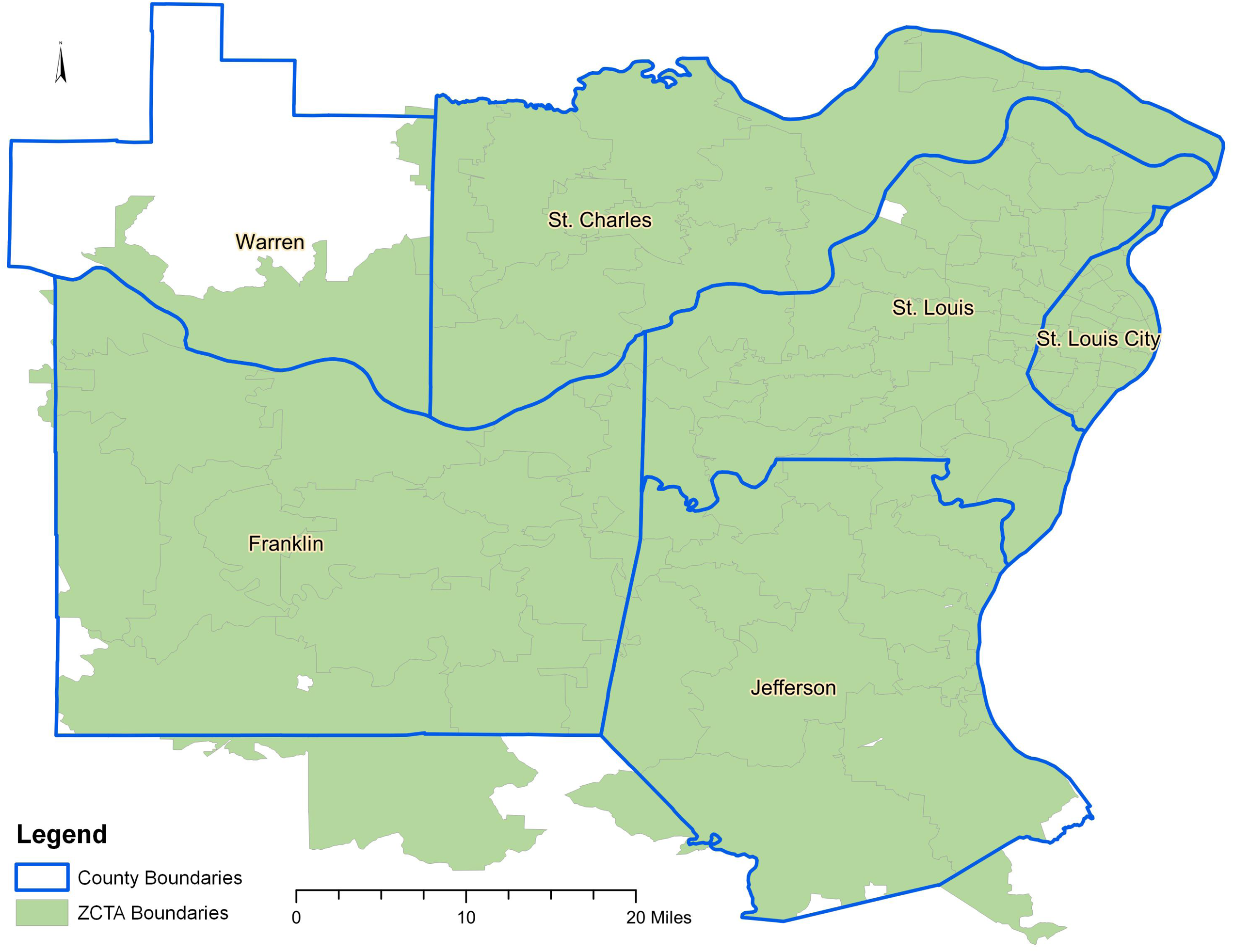
Map of study area showing geographic distribution of Zip Code Tabulation Areas and Counties.

### Data Sources

#### COVID-19 and Chronic Disease Hospitalization Data

Data on COVID-19 and chronic condition hospitalizations were obtained from the Hospital Industry Data Institute, a not-for-profit organization founded by the Missouri Hospital Association that summarizes state level discharge data. The COVID-19 data provided information on the numbers of COVID-19 discharges by patient ZCTA and by age group from April 1, 2020 to September 30, 2020. In addition to the COVID-19 hospitalization data, data on the number of confirmed COVID-19 cases were obtained from the county Departments of Health. COVID-19 case data were included as one of the potential predictors of COVID-19 hospitalization risks. Data on chronic conditions were extracted for the time period 2019-2020 based on ICD-10 codes and included the following conditions/behaviors: obesity, tobacco use, cancer (breast, colorectal, prostate, lung, endometrial, leukemia and lymphoma), chronic obstructive pulmonary disease, chronic kidney disease, heart failure, and diabetes. These conditions were selected due to their potential association with COVID-19 severity. All data were aggregated to the ZCTA-level to facilitate subsequent ZCTA-level analyses. The ZCTA-level risks of these conditions were then computed and presented as disease-specific hospitalizations per 100 population.

#### Socioeconomic, demographic and base map data

ZCTA-level data on socioeconomic and demographic factors including age, sex, race, population density, education level, and median income, were obtained from the U.S. Census Bureau 2018 American Community Survey (ACS) 5-year estimates [15]. The cartographic boundary files, used for generating maps, were obtained from the US Census Bureau Website [16].

### Descriptive Analyses

All descriptive analyses were performed in R version 4.1.0 [17] using RStudio version 1.4.1103 [18]. Normality of distribution of continuous variables was assessed using Shapiro-Wilk test. Medians as well as 1^st^ and 3^rd^ quartiles were used as measures of central tendency and spread for all continuous variables since they were all non-normally distributed. The COVID-19 ZCTA-level hospitalization risks were directly age-standardized, in STATA version 16 [19], using the 2010 US population as the standard population.

### Investigation of Predictors of COVID-19 Hospitalizations

#### Global Models

Univariable global Poisson models were used to assess simple associations between each of the potential predictors and COVID-19 hospitalization risk. As opposed to local models that estimate as many regression coefficients per predictor as the number of geographic units, global models assume constant relationships between each potential predictor and the outcome and therefore estimate one regression coefficient per potential predictor. These models were fit under the Generalized Linear Model (GLM) framework in R [17] specifying log link. The dependent variable was the expected count of COVID-19 hospitalizations in each ZCTA based on the age-adjusted/standardized number of COVID-19 hospitalizations and the offset was the natural log of the population in each ZCTA. A relaxed α of 0.15 was used to assess potentially significant predictors in univariable models. The linearity of the relationships between the log risk and potential predictor variables was assessed graphically.

Spearman rank correlation coefficients were computed to assess bivariate correlations among the potential predictor variables identified during the univariable analyses using R [17]. Variables with correlation coefficients greater than 0.7 were considered highly correlated. To avoid multicollinearity, only one of a pair of highly correlated variables was retained for further assessments using multivariable Poisson model. Biological and statistical considerations were used to determine which of a pair of highly correlated variables to retain. Non-correlated variables that had univariable p≤0.15 were assessed in a multivariable global Poisson model, built using manual backwards elimination and an α of 0.05. Variables were considered as confounders if their removal from the model resulted in changes of at least 20% of the coefficients of any of the variables in the model. Confounders were retained in the final main effects model. Biologically plausible two-way interactions between variables in the final main effects multivariable model were assessed and the significant ones retained in the final model. The final Poisson regression model was then assessed for overdispersion.

Due to the presence of overdispersion in the final Poisson model (based on comparison of model deviance and degrees of freedom), a global negative binomial model was built. The process of building the negative binomial (NB) model was similar to that of the Poisson model described above. However, the glm.nb function of the MASS package [20] of R [17] was used instead of the glm function. The rest of the model specifications were similar to the Poisson model. Goodness-of-fit of the final global NB model was assessed using Pearson and Deviance chi-square goodness-of-fit tests.

#### Local Models

To assess if the association between each of the predictors and hospitalization risks varied by geographic location, a geographically weighted negative binomial (GWNB) model, proposed by Silva and Rodrigues [21], was fit to the data specifying the same outcome, link function, offset and significant predictors as in the final global multivariable NB model. However, unlike the global multivariable NB model that estimates one regression coefficient for each predictor and thus assumes a constant strength of association across all ZCTAs, the GWNB theoretically estimates as many regression coefficients as the number of ZCTAs. Essentially, GWNB evaluates a local model of the outcome by fitting a regression equation to every ZCTA in the dataset.

These separate model equations are constructed by incorporating the outcome and predictor variables of the ZCTAs that fall within the neighborhood of each target ZCTA. Therefore, the GWNB allows identification of local variations in the strength of associations and therefore giving the importance of specific predictors in different local areas (ZCTAs). This implies that some factors may be more important predictors of hospitalization risk in some ZCTAs than others.

The local GWNB model was fit in SAS version 9.4 [22] using a SAS/IML macro [23]. The estimation of local regression coefficients was based on biquadratic kernel weighting function [23], while the bandwidth was estimated using the adaptive method which allows the size of the bandwidth to vary based on the density of observations. Bias-corrected Akaike Information Criteria (AICc) was used to determine the optimum kernel bandwidth and for comparing the goodness-of-fit of the global NB and GWNB models. The better fitting model was identified as the one with the lower AICc value.

Stationarity of the GWNB coefficients was assessed using: (a) randomization non-stationarity test based on 999 replications [24]; (b) comparison of the interquartile range of the local GWNB model coefficients with the standard error estimates of the global NB model. Local coefficients whose interquartile ranges were larger than twice the standard error of the regression coefficient from the global NB model were considered non-stationary [25, 26].

### Cartographic Displays

Choropleth maps showing the geographic distributions of ZCTA-level age-adjusted COVID-19 hospitalization risks, the socioeconomic, demographic and chronic disease factors as well as local regression coefficients from the GWNB models were generated using QGIS 3.16.6 [27]. Jenk’s optimization classification scheme [28–30] was used to determine the critical intervals of the choropleth maps.

## Results

### Descriptive Statistics

The ZCTA-level median percentage of males was 48.6% while that of black and Hispanic populations were 3.7% and 2.2%, respectively (**Table 1**). For education variables, 38.2% of the population had high school education, 23% had some college education while 8.6% had associate’s degree and 18% had bachelor’s degree. The median household income was just over $59,400 with 9.5% of the ZCTA-level population living below the poverty line. Among chronic conditions investigated, Chronic Kidney Disease had the highest hospitalization risk (7.6%) followed by diabetes (7.5%) and heart failure had the lowest (3.2%) (**Table 1**). The ZCTA-level median number of confirmed cases of COVID-19 was 360 cases which was equivalent to 2.2% of the population at ZCTA-level (**Table1**).

**Table 1:** Descriptive Statistics of ZCTA-level Potential Predictors of COVID-19 Hospitalization Risks in the St. Louis Area, Missouri.

| Type of Variable | Variable | Median | First Quartile | Third Quartile |
| --- | --- | --- | --- | --- |
| Demographic Factors |  |  |  |  |
|  | % male population | 48.6 | 47.4 | 50.3 |
|  | % black population | 3.7 | 0.9 | 34.2 |
|  | % Hispanic/Latino population | 2.2 | 1.0 | 3.2 |
| Educational Variables |  |  |  |  |
|  | % with ≤ high school education | 38.2 | 23.9 | 47.6 |
|  | % with some college | 23.0 | 19.8 | 25.6 |
|  | % with associate's degree | 8.6 | 6.6 | 10.6 |
|  | % with bachelor's degree | 18.0 | 9.4 | 25.9 |
| Economic Variables |  |  |  |  |
|  | % below poverty level | 9.5 | 5.8 | 16.8 |
|  | median household income | 59,768.5 | 46005.3 | 77504.0 |
| Health Behavior (ZCTA-level number of hospitalized patients that use tobacco per 100) |  |  |  |  |
|  | % tobacco<sup>1</sup> | 10.4 | 6.8 | 14.5 |
| Co-morbidities (ZCTA-level number of hospitalized patients with specific condition per 100) |  |  |  |  |
|  | % obesity | 7.0 | 5.6 | 8.0 |
|  | % diabetes | 7.5 | 6.5 | 9.2 |
|  | % cancer | 3.8 | 3.3 | 4.3 |
|  | % COPD<sup>2</sup> | 4.0 | 3.1 | 4.9 |
|  | % CKD<sup>3</sup> | 7.6 | 6.4 | 8.9 |
|  | % heart failure | 3.2 | 2.6 | 3.8 |
| COVID-19 Cases |  |  |  |  |
|  | total cases | 360 | 118 | 607 |
|  | cases per 100 population | 2.2 | 1.9 | 2.5 |

<sup>1</sup>ZCTA-level % of COVID-19 hospitalized patients that were tobacco users
<sup>2</sup>Chronic Obstructive Pulmonary Disease
<sup>3</sup>Chronic Kidney Disease

### Predictors of COVID-19 Hospitalization Risks

#### Global Model

A number of variables had univariable associations with COVID-19 hospitalization risks at a relaxed p<0.2 (**Table 2**). Of the assessed demographic variables, only percentage of black population had a univariable association (p<0.001) with hospitalization risk. By contrast, all the assessed educational, economic, health behavior and chronic disease variables had significant univariable associations with COVID-19 hospitalization risks (**Table 2**). The ZCTA-level risk of confirmed COVID-19 cases had a significant (p<0.001) univariable association with COVID-19 hospitalization risk but the raw count of COVID-19 cases per ZCTA did not (p=0.9) (**Table 2**).

**Table 2:** Univariable Associations of Sociodemographic, Economic, and Chronic Disease Potential Predictors of COVID-19 Hospitalization Risk in the St. Louis Area, Missouri.

| Type of Variable | Variable | Coefficient | 95% Confidence Interval | P-values |
| --- | --- | --- | --- | --- |
| Demographic Factors |  |  |  |  |
|  | % male population | 0.004 | -0.015, 0.024 | 0.727 |
|  | % black population | 0.007 | 0.005, 0.009 | <0.0001 |
|  | % Hispanic/Latino population | -0.021 | -0.056, 0.016 | 0.292 |
| Educational Variables |  |  |  |  |
|  | % with ≤ high school education | 0.015 | 0.010, 0.020 | <0.0001 |
|  | % with some college education | 0.035 | 0.022, 0.049 | <0.0001 |
|  | % with associate's degree | 0.028 | 0.0002, 0.055 | 0.036 |
|  | % with bachelor's degree | -0.021 | -0.028, -0.014 | <0.0001 |
| Economic Variables |  |  |  |  |
|  | % below poverty level | 0.017 | 0.009, 0.024 | <0.0001 |
|  | median income | -5.00E-06 | -0.000008, -0.000003 | <0.0001 |
| Health Behavior (ZCTA-level number of hospitalized patients that use tobacco per 100 Population) |  |  |  |  |
|  | % tobacco<sup>1</sup> | 0.046 | 0.034, 0.058 | <0.0001 |
| Co-morbidities (ZCTA-level number of hospitalized patients with specific condition per 100 Population) |  |  |  |  |
|  | % obesity | 0.138 | 0.104, 0.171 | <0.0001 |
|  | % cancer | -0.029 | -0.096, 0.032 | 0.377 |
|  | % COPD<sup>2</sup> | 0.11 | 0.055, 0.164 | <0.0001 |
|  | % CKD<sup>3</sup> | 0.1 | 0.068, 0.131 | <0.0001 |
|  | % heart failure | 0.194 | 0.123, 0.265 | <0.0001 |
|  | % diabetes | 0.11 | 0.083, 0.136 | <0.0001 |
| Confirmed COVID-19 Cases |  |  |  |  |
|  | total cases | 1.40E-05 | -0.0002, 0.00023 | 0.9 |
|  | cases per 100 population | 0.277 | 0.192, 0.3621 | <0.0001 |

<sup>1</sup>ZCTA-level % of COVID-19 hospitalized patients that were tobacco users
<sup>2</sup>Chronic Obstructive Pulmonary Disease
<sup>3</sup>Chronic Kidney Disease

Based on the final global multivariable NB model, the ZCTA-level hospitalization risks were higher in ZCTAs that were high in the following predictors: percentage of black population (p=0.0416), percentage of population with some college education (p=0.0005), percentage of individuals hospitalized with diabetes (p<0.0001), and number of ZCTA-level COVID-19 cases per 100 population (p=0.0001) (**Table 3**). A map of the distribution of age-adjusted COVID-19 hospitalization risks and each of the significant predictors is shown in **Figure 2**. High age-adjusted COVID-19 hospitalization risks tended to occur in the Northeast of the study area and included ZCTAs in parts of St. Charles, St. Louis and Louis City counties. These areas also had high percentages of black population, individuals with some college education, those with high diabetes hospitalization risks as well as high risks of COVID-19 cases (**Figure 2**). High hospitalization risks were also evident in the Southwest areas of the study area that included parts of Warren and Franklin counties. It is worth noting that these areas also tended to have high diabetes hospitalization risks and percentages of the population with some college education (**Figure 2**). Since the global model did not show evidence of good fit based on both Deviance (p=0.03) and Pearson (p=0.01) goodness-of-fit tests, stationarity of the regression coefficients was assessed using a local GWNB model.

**Table 3:** Final Global Negative Binomial Model Showing Significant Determinants of COVID-19 Hospitalization Risk in the St. Louis area.

| Name | Coefficient | 95 % Confidence Interval | p-value |
| --- | --- | --- | --- |
| % black population | 0.0014 | 0.0001, 0.0027 | 0.0416 |
| % with some college education | 0.0180 | 0.0078, 0.0281 | 0.0005 |
| % diabetes<sup>1</sup> | 0.0628 | 0.0397, 0.0860 | <0.0001 |
| Confirmed COVID-19 cases per 100 population | 0.2623 | 0.2027, 0.3218 | <0.0001 |

<sup>1</sup>Number of hospitalized patients with diabetes per 100 Population

**Figure 2:**
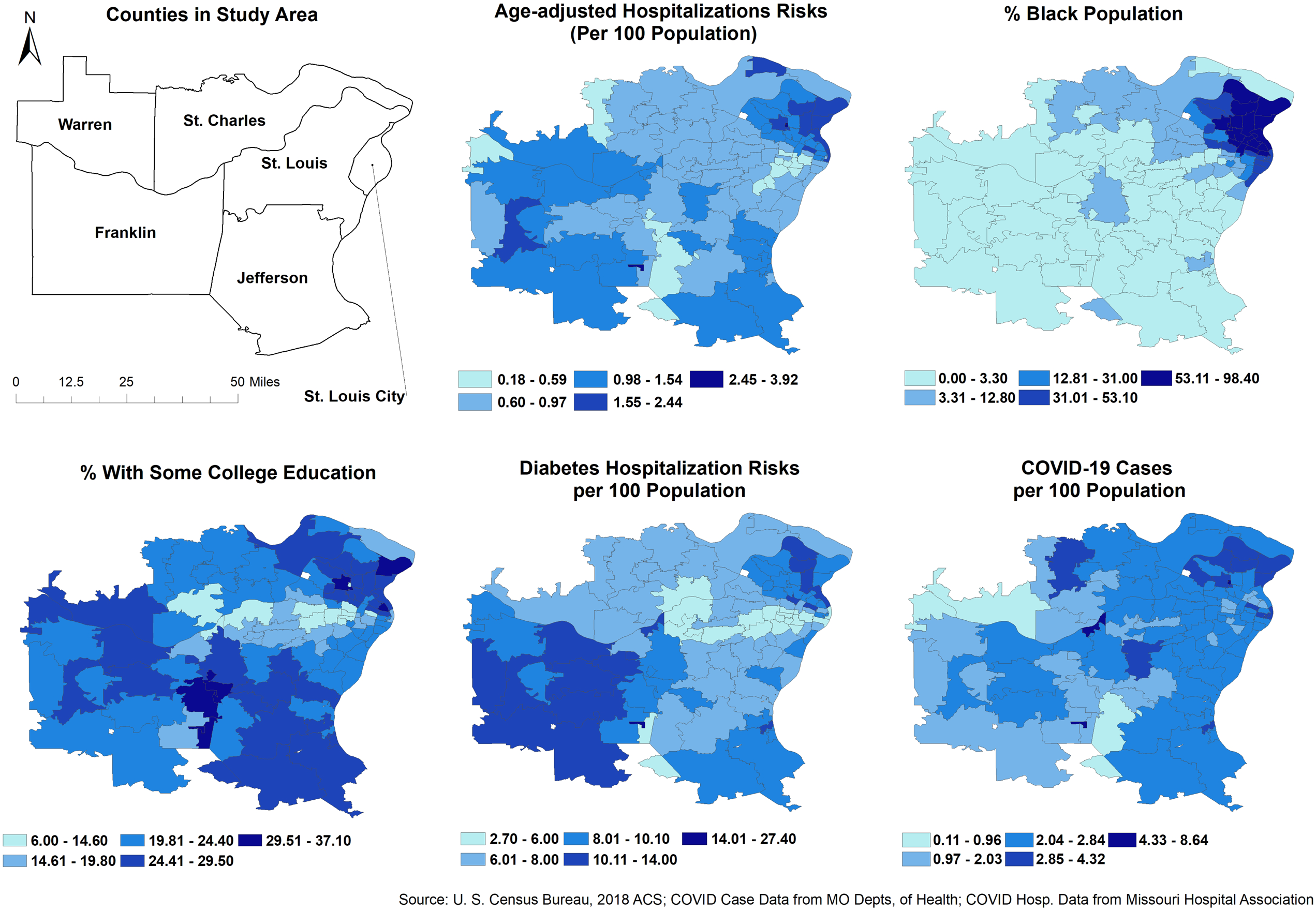
Geographic distribution of ZCTA-level COVID-19 age-adjusted hospitalization risks and its significant predictors in the St. Louis area, Missouri.

#### Local Model

The p-values of the stationarity tests from the GWNB model indicate that the coefficients for the association between COVID-19 hospitalization risks and percentage of black population (p=0.001) and number of hospitalized patients with diabetes per 100 population (p=0.032) were non-stationary (**Table 4**). Additionally, comparison of the 2×SEs of the global coefficients and IQR of the local coefficients showed evidence of non-stationarity of the coefficients of the above two predictors as well as the population adjusted cases of COVID-19 (**Table 4**).

**Table 4:** Results of assessment of stationarity of the coefficients of the predictors of the COVID-19 hospitalization risks in the St. Louis Area, Missouri.

|  | Global<br>SE<sup>1</sup> | Global<br>SE<sup>1</sup>x2 | IQR<sup>2</sup> of Local<br>Coefficient | IQR-2(SE) | Stationarity<br>test p-value | Is Coefficient<br>Stationary? |
| --- | --- | --- | --- | --- | --- | --- |
| % Black Population | 0.0007 | 0.0014 | 0.0148676 | 0.0134676 | 0.001 | No<sup>4</sup> |
| % With Some College Education | 0.0052 | 0.0104 | 0.0092401 | -0.0011599 | 0.406 | Yes |
| % diabetes<sup>3</sup> | 0.0118 | 0.0236 | 0.0416903 | 0.0180903 | 0.032 | No<sup>4</sup> |
| Cases of COVID-19 per 100 population | 0.0304 | 0.0608 | 0.1164299 | 0.0556299 | 0.109 | No<sup>5</sup> |

<sup>1</sup>Standard Error
<sup>2</sup>Interquartile Range
<sup>3</sup>Number of hospitalized patients with diabetes per 100 Population
<sup>4</sup>Coefficients are non-stationary based on both the p-value of the stationarity test and IQR-2(SE) assessment
<sup>5</sup>Coefficient is non-stationary based on IQR-2(SE) assessment

The spatial distribution of the local coefficients of the three predictors whose relationship with COVID-19 hospitalizations were identified as non-stationary provides visual evidence for variability of the local relationships (**Figure 3**). Thus, associations of COVID-19 hospitalization risks with percentage of black population, diabetes hospitalization risks and COVID-19 adjusted cases varied considerably across the study area, with a strong East-West gradient. The association between percentage of black population and COVID-19 hospitalization risks, for instance, was positive in the Northeast and negative in the West and Southwest. Moreover, the strength of the association was higher in ZCTAs in the West compared to those near the center of the study area. The strength of association between COVID-19 hospitalization risks with diabetes hospitalization risks was also higher in the Northeast and lower in the West. All ZCTAs, except one (63025), showed evidence of positive association between COVID-19 hospitalization risks and diabetes hospitalization risks. Finally, the association between COVID-19 hospitalization and population adjusted cases of COVID-19 increased from West to East, with the association staying positive in all ZCTAs of the study area. It is worth noting that the local GWNB model had a much better goodness- of-fit (AICc=986.4) than the global model (AICc=1002.7).

**Figure 3:**
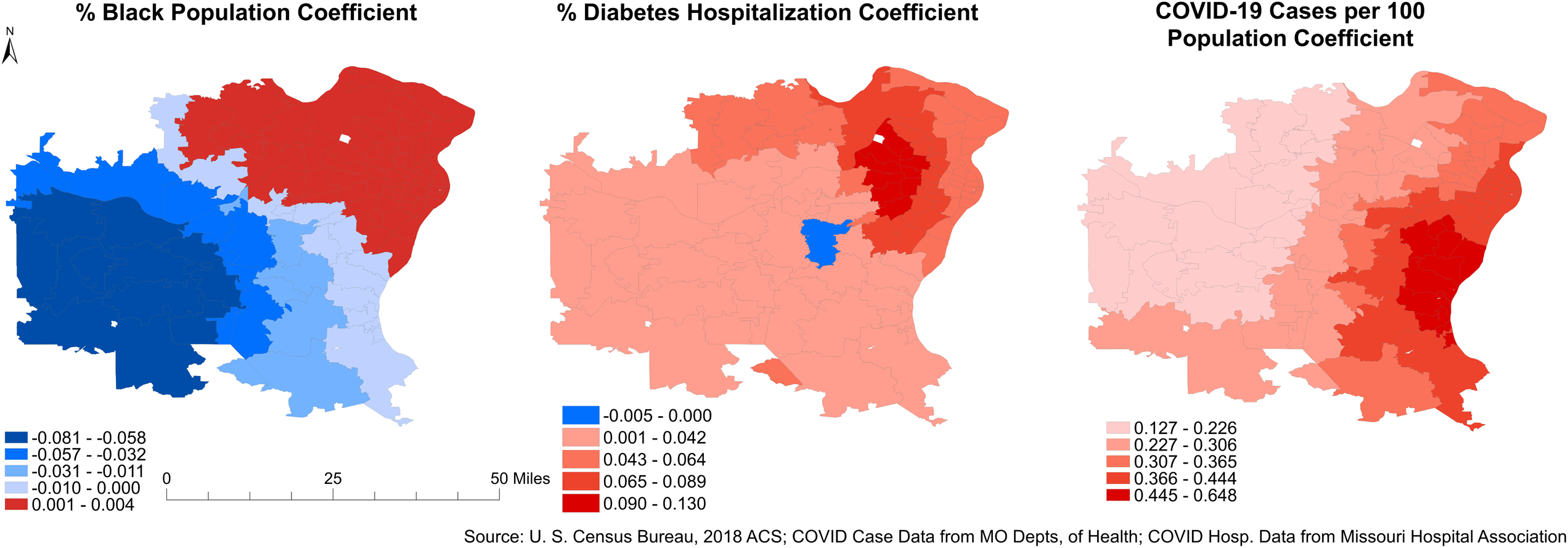
Geographically Varying Coefficients of the Local Geographically Weighted Negative Binomial Model Predicting COVID-19 Age-adjusted Hospitalization Risks in the St. Louis area.

## Discussion

The goal of this study was to investigate geographic disparities and identify predictors of ZCTA-level COVID-19 hospitalization risks in the St. Louis area. The findings of this study can be used to identify areas where the population is at higher risk of hospitalization due to COVID-19 in order to guide planning and control efforts and to reduce potential overburdening of hospitals during COVID-19 surges.

There was evidence of geographic disparities in COVID-19 age-adjusted hospitalization risks in the study area. Urban ZCTAs in St. Louis City and St. Louis County exhibited high hospitalization risks. These ZCTAs also have high percentages of the population that are black, some as high as 98.4%. Some of these ZCTA also had high diabetes hospitalization risks. It is worth noting that some rural ZCTAs in Franklin and Warren counties had high COVID-19 hospitalization risk but very low percentages of the population that were black. However, these ZCTAs had high diabetes hospitalization risks implying that the COVID hospitalization risks in these rural ZCTAs was more driven by diabetes burden than demographic factors. Thus, although COVID-19 hospitalization risks in the more urban areas seems to be driven by the demographic composition of the population, the risks in the more rural areas seem to be driven more by diabetes burden. Geographic areas with intermediate to high COVID-19 hospitalization risks tended to have high percentages of the population with some college education and included ZCTAs in Franklin, Jefferson, St. Louis City and St. Louis County. It is worth noting that St. Louis City and St. Louis County tend to have similar restrictions, but often time, the restrictions from the other counties are more laxed. Previous ecological and individual level studies have not considered education level as a predictor of hospitalization risk. The level of education may be a proxy of occupation and other sociodemographic factors that may impact both the risk of infection and the resulting severity of the disease and hence hospitalization risks.

The above findings are consistent with those from previous ecological studies that investigated risk factors and predictors of COVID-19 hospitalization. For instance, an ecological study by Nguyen et al. reported that diabetes was a significant predictor of increased COVID-19 hospitalization risk at the county level in Georgia, USA even after controlling for sociodemographic and economic factors [31]. On the contrary, an Iranian study conducted at the provincial level did not find a significant association when only controlling for chronic disease factors [32]. This may indicate differences of relationships in different geographic areas and populations as well as the importance of controlling for sociodemographic factors when evaluating the impact of chronic disease variables on COVID-19 hospitalization risk. Considering the fact that ZCTAs with high diabetes hospitalization risks also tended to have high COVID-19 hospitalization risks, COVID-19 mitigation efforts may need to be targeted to these ZCTAs to reduce the potential burden of the disease.

Interestingly, the study by Nguyen et al., which also considered sociodemographic and economic factors as well as comorbidities, did not find a significant association between percentage of black population with COVID-19 hospitalization risk [31]. However, it did identify other socioeconomic factors that this study did not consider, including: percentage of children in poverty and percentage of the population with severe housing problems. Although not directly comparable with the current ecological study, previous individual level studies have also identified associations between black race and COVID-19 hospitalization risk [8, 13, 33].

The local GWNB model allowed modeling of geographically varying associations between COVID-19 hospitalization risks and its predictors instead of assuming constant associations across the study area. Although the coefficients of percentage of black population, diabetes hospitalization risk and risk of COVID-19 infections varied spatially, the coefficients of percentage of the population with some college education did not and hence was modeled as stationary. This suggests that the coefficients for the percentage of the population with some college education are generalizable to all ZCTAs in the study area. In contrast, geographic variations in the associations between COVID-19 hospitalization risks and the percentage of black population, diabetes hospitalization risks and risks of COVID-19 infections suggest that global coefficients do not adequately describe the associations between COVID-19 hospitalization risks and these predictors across the study area. These findings have health planning and service provision implications. For instance, a “one size fits all” approach would not be suitable for addressing geographic disparities in COVID-19 hospitalization risks across the study area since some predictors are more important in some locales than others. Thus, different locales may require slightly different strategies depending on the most important predictors driving COVID-19 hospitalization risk in the location. Therefore, planning for hospital capacity and other disease management and control efforts will need to use evidence-based approaches informed by empirical evidence from both global and local models.

### Strengths and limitations

This study used both global and local models to investigate geographic disparities and identify predictors of COVID-19 hospitalization risks in St. Louis region of Missouri. The use of local models to investigate stationarity of regression coefficients of significant predictors and model non-stationary coefficients is a key strength of the study. This approach is particularly important in guiding local health planning since the importance of different predictors are not constant across the study area implying that different management and control strategies may need to be used in different areas. Therefore, modeling approaches that use both global and local models help to better understand the relationships between the outcome and predictors and may be more useful in guiding control efforts at the local level. However, the study is not without limitations.

The hospital data has limitations associated with diagnostic classifications of COVID-19 in situations when the patient has co-morbidities that may have contributed to hospitalization. Additionally, there may be geographic differences in COVID-19 case ascertainment and reporting.

## Conclusions

There is evidence of geographic disparities in COVID-19 hospitalization risks in the St. Louis area of Missouri. These disparities are driven by socioeconomic, demographic and health-related factors. The impacts of these factors vary by geographical location with some factors being more important predictors of COVID-19 hospitalization risk in some locales than others. This demonstrates the importance of using not only global but also local models to investigate determinants of geographic disparities in health outcomes and utilization of health services. This study’s findings are useful for informing healthcare system planning to identify geographic areas likely to have high numbers of individuals needing hospitalization as well as in guiding vaccination efforts.

## Data Availability

The investigators cannot share the datasets, which are not publicly available, because they do not have legal ownership nor authority to share the data. However, the data are available upon request from the Hospital Industry Data Institute

## Declarations

### Ethical approval and consent to participate

The study was approved by the North Carolina State University Institutional Review Board (IRB number: 22342) and all study methods were carried out in accordance with relevant guidelines and regulations. The study used anonymized secondary data provided to the investigators, by the Hospital Industry Data Institute, in such a manner that the identify of human subjects cannot be ascertained directly or through identifiers linked to the subjects. The investigators did not contact the subjects and did not re-identify subjects.

### Consent for publication

Not applicable.

### Availability of data and materials

The investigators cannot share the datasets, which are not publicly available, because they do not have legal ownership nor authority to share the data. However, the data are available upon request from the Hospital Industry Data Institute at 4712 Country Club Drive P.O. Box 60 Jefferson City, MO 65102-0060, telephone (573)893-3700.

### Authors’ contributions

SL, ALL, CL, and AO conceptualized research idea

LL, DT collected and curated the data

MI, PD, SL, ALL, CL and AO analyzed data

MI, PD and AO wrote the manuscript

All authors edited the manuscript

All authors read and approved the final manuscript.

## Acknowledgements

We thank the St. Louis Comparative Modeling Network for facilitating access to data and useful discussions.

